# Single Dose Pharmacokinetic Comparison of Citrulline Dietary Supplements

**DOI:** 10.1101/2025.09.19.25332543

**Authors:** Jon C Wagner, William Faulkner, Freddy Mullins, Mark Faulkner

## Abstract

Citrulline dietary supplements are of interest for improving cardiovascular and exercise performance. The health benefits of citrulline are attributable to its ability to increase arginine levels in the body through the intestinal-renal arginine conversion pathway. While L-citrulline is the most widely used citrulline dietary supplement, additional salt forms such as citrulline maleate and citrulline HCl may provide improvements in the delivery of arginine to the body. The present study compared the single-dose pharmacokinetics of L-citrulline and citrulline HCl formulations in healthy human subjects. A total of 17 subjects were randomly assigned to groups receiving either a standard 6 g dose of L-citrulline, or a 6 g or 2 g dose of citrulline HCl. Measurement of citrulline and arginine in plasma and urine samples were determined using liquid chromatography-multiple reaction monitoring mass spectrometry (LC-MRM/MS). All citrulline dietary supplements examined produced time-dependent increases in plasma citrulline and arginine. Despite lower citrulline maximal plasma concentration (Cmax) and area under the plasma vs time curve (AUC) in subjects receiving 6 g citrulline HCl, the resulting arginine Cmax and AUCs were similar to that observed with 6 g L-citrulline. Subjects receiving the 2 g dose of citrulline HCl had lower Cmax and AUC values compared to 6 g doses, however, the time required to reach peak levels of arginine occurred in half the time required for the L-citrulline group (60 minutes for citrulline HCl vs 120 minutes for L-citrulline). While all supplements resulted in increased urinary excretion of citrulline, the elevations were greatest for L-citrulline (45-fold) compared to the 6g (15-fold, 3x less than L-citrulline) and 2g (7-fold, >6x less than L-citrulline) citrulline HCl formulations. The decreased urinary elimination with citrulline HCl is consistent with the improved citrulline to arginine conversion rate observed with the citrulline HCl formulations, and likely accounts for the increased relative arginine bioavailability of the 6 g citrulline HCl (125%) and 2 g citrulline HCl (226%), compared to L-citrulline. Together these studies indicate that dietary supplementation with 2 g citrulline HCl provided a faster and more efficient method for increasing plasma arginine than the L-citrulline formulation.

## Introduction

Citrulline is a non-essential amino acid that plays an important role in nitrogen homeostasis. It is an intermediate metabolite in the urea cycle preventing the build-up of toxic levels of ammonia in the body (1). Citrulline is also involved in arginine and nitric oxide (NO) production which has important cardiovascular functions in the regulation of blood flow and vasodilation (1-3). Citrulline can be obtained through the diet, with the consumption of watermelon, cucumbers, pumpkin and squash providing a rich source of citrulline. Citrulline is also synthesized from arginine and glutamine within the intestinal enterocytes (1, 2). Unlike arginine which has limited intestinal permeability and undergoes significant first-pass metabolism in the liver, citrulline is readily absorbed from the gastrointestinal system and the resulting circulating plasma citrulline undergoes conversion to arginine within the kidney which is then available for localized NO production within the vasculature (Figure 1). Indeed, this intestinal-renal axis for arginine synthesis makes dietary supplementation with citrulline a highly effective method for enriching arginine levels in the body that are important for cardiovascular response, protein production and muscle performance.

**Figure 1.**
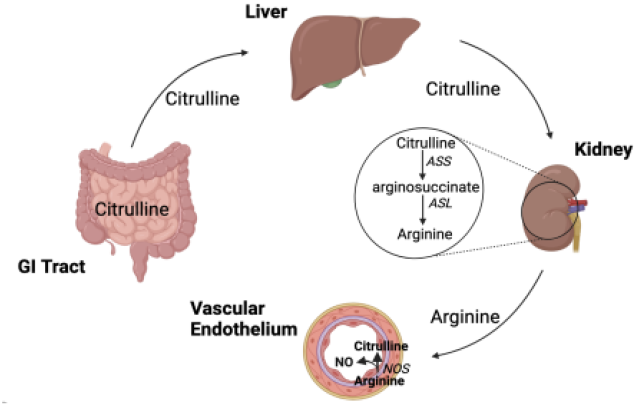
Dietary citrulline as a source of arginine for the body. Citrulline in the gastrointestinal tract is readily absorbed and avoids first-pass metabolism in the liver to enter into systemic bloodstream. Citrulline in the systemic bloodstream is converted into arginine through a two-step enzymatic process in kidney involving arginosuccinate synthetase (*ASS*) and arginosuccinate lyase (*ASL*). Arginine formed in the kidney enters into systemic circulation where it is available for uptake into tissue for protein synthesis. Arginine taken up by vascular endothelial cells is also available for conversion to NO via nitric oxide synthase (*NOS*).

Several studies have demonstrated dietary supplementation with L-citrulline effectively increases arginine levels in healthy individuals (4-6). From a health perspective, dietary supplementation with L-citrulline has shown a variety of beneficial effects including restoring microvascular perfusion during endotoxemia (7), improving exercise performance and lowering blood pressure in post-menopausal women (8, 9), preventing postoperative pulmonary hypertension in pediatric patients undergoing cardiopulmonary bypass surgery (10). While daily doses of L-citrulline of up to 10 g have been used without GI distress, most studies use 6 g or less (4).

The primary form of citrulline used in dietary supplementation is L-citrulline. Other salt forms of citrulline, including citrulline malate, have been reported to have improved effects (11, 12), although other studies have failed to show significant differences over L-citrulline (13). Citrulline HCl is a new salt form with improved aqueous solubility over L-citrulline. Given the standard dosages for most citrulline dietary supplements (6 g), the improved aqueous solubility of the hydrochloride salt may provide improved intestinal absorption and a more favorable gastro-intestinal side-effect profile than other citrulline formulations. The present study compared the pharmacokinetics of a standard 6 g daily dose of L-citrulline with either a 6 g or 2 g dose of citrulline HCl. Plasma levels of both citrulline and arginine were increased with all supplements examined, however, total arginine conversion was greater with citrulline HCl compared to L-citrulline. Compared to L-citrulline, both the 6 g and 2 g doses of citrulline HCl salt resulted in higher relative arginine bioavailability, 125% (6 g HCl) and 225% (2 g HCl) compared to the L-citrulline supplementation group.

## 2. Methods

### 2.1 Study design

The study (IRB2024-SP100) was approved by the UNIVO investigational review board on June 30, 2024 and registered on clinicaltrials.gov (NCT06977854). Recruitment, enrollment, and performance of the study was conducted by Princeton Consumer Research (Raritan, NJ). Healthy adult men and women enrolled in the study were randomly assigned to one of the following treatment arms; 6 g L-citrulline (L-Cit), 6 g Citrulline HCl (Cit HCl), or 2 g Cit HCl. On the testing day, subjects having fasted at least 3-hours were administered the various dietary supplements mixed in 125 ml of water. Venous blood samples were collected in heparinized tubes prior to treatment and at 30, 60, 90, 120, 150 and 180-minutes following treatment. Twenty-four-hour urine collection was done prior to citrulline supplementation and immediately following. Blood and urine samples were centrifuged, and the resulting plasma and urine was stored at -80° C until analyzed as described below.

### 2.2 Sample Analysis

The plasma and urine samples were analyzed for citrulline, arginine, and creatinine using liquid chromatography-multiple reaction monitoring mass spectrometry (LC-MRM/MS) on an Agilent 1290 UHPLC system connected to an Agilent 6495B triple-quadrupole mass spectrometer at the University of Victoria Analytical Laboratory. Briefly, an internal standard (IS) solution consisting of arginine-^13^C_6_, argininic-d_6_ acid, creatinine-d_3_ and citrulline-d_7_ was prepared in a 6% trichloroacetic acid solution. Plasma (50 μl) and urine (100 μl) samples were mixed with 100 μl of IS and the mixtures were vortexed for 10 s at 3,000 rpm and then sonicated in an ice-water bath for 1-min, followed by centrifugal clarification at 21,000 g and 10 °C for 10 min. Plasma (50 μl) and urine (100 μl) samples were mixed 1:4 and 1:10, respectively, with de-ionized distilled water. A 4 μl aliquot of the resulting samples and calibration solutions were analyzed using LC-MRM/MS. A Waters HSS T3 UPLC column (2.1*100 mm, 1.8 μm) was used for the chromatographic separation of analytes with a 5-mM heptafluorobutyric acid acetonitrile mobile phase for binary-solvent gradient elution under optimized conditions. The LC-MRM/MS raw data files were acquired and processed using the Agilent MassHunter® software suite. For concentration calculations, linearly regressed calibration curves of the analytes were constructed with the data acquired from the calibration solutions. Concentrations of the analytes in plasma and urine samples were calculated by interpolating the calibration curve with the peak area ratios areas measured from the sample solutions. Both samples and calibration solutions were run in duplicate.

### 2.3 Pharmacokinetic Calculations

The baseline and maximum concentration of arginine and citrulline in plasma (Cmax), time required to reach peak levels (Tmax), and area under the plasma concentration vs time curve (AUC) were obtained for each individual subject. The AUC_(0-180min)_ for arginine and citrulline were determined using the linear trapezoidal method. Estimation of the AUC_(0-∞)_ was done using the following formula: AUC(_0-180min)_ + C_180_ /λ_z_; where C_180_ is the plasma concentration at 180 minutes and λ_z_ is the elimination rate constant obtained by linear regression analysis of the terminal phase of the plasma concentration (log scale) vs time curve. The AUC_(0-∞)_ ratios for plasma arginine and plasma citrulline were used to estimate the fraction of citrulline converted to arginine and provide an index of the conversion efficiency for each of the citrulline dietary supplements examined. The relative bioavailability of the citrulline HCl formulations compared to the standard L-citrulline formulation were determined for both citrulline and arginine using the following formula: 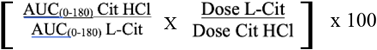

The creatinine clearance was calculated using (Urine Creatinine Concentration x 24-hr urine volume) / (Plasma Creatinine Concentration x time). Renal clearance of arginine and citrulline were determined in a similar fashion using urine concentrations from the 24-hr urine samples. The plasma AUC_(0-∞)_/24 were used to obtain an average plasma concentration for arginine and citrulline over the 24-hour period of urine collection. The fractional reabsorption rate (Fr) for arginine and citrulline was calculated as previously described (4) using the following equation: 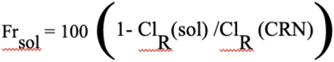

Where Fr_sol_ is the fractional reabsorption of the solute (either arginine or citrulline) and Cl_R_ (sol) is the renal clearance of the solute and Cl_R_ (CRN) represents the renal clearance of creatinine.

### 2.4 Statistical Analysis

Data are expressed as the mean ± standard deviation unless otherwise stated. Graphs were constructed using Excel and Graph Robot (14). Statistical analysis of mean values between treatment groups were performed using one way ANOVA followed by Fisher LSD test of the individual means.

## 3. Results

### 3.1 Study Demographics

All of the 23 subjects enrolled in the study completed the study without any adverse events reported. As kidney function is critical for both the conversion of citrulline to arginine and the reabsorption of amino acids from the urine, 24-hr creatinine clearance was determined as a measure of kidney function. A total of 6 subjects (2 in each treatment group) were excluded due to 24-hr creatinine clearance of less than 60 ml/min. The demographics of the remaining subjects in the various citrulline supplementation groups is shown in Table 1.

**Table 1:** Subject demographics and renal function assessment.

| Treatment Group | Amount of citrulline per dose | # Males / Females (% Female) | Age | | CRN CL<sub>R</sub> (ml/min)<br>Avg ± SD |
| --- | --- | --- | --- | --- | --- |
| | | | Avg ± SD | Range | |
| L-citrulline - 6 g (L-Cit 6g) | 6.0 g (34 mmole) | 2 / 4 (67%) | 48 ± 12 | 31 - 59 | 99 ± 33 |
| Citrulline HCl – 6 g (Cit HCl 6g) | 4.9 g (28 mmole) | 2 / 4 (67%) | 46 ± 14 | 31 - 62 | 80 ± 24 |
| Citrulline HCl – 2 g (Cit HCl 2g) | 1.7 g (8 mmole) | 2 / 3 (60%) | 44 ± 15 | 29 - 61 | 85 ± 25 |

### 3.2 Elevations in Plasma Citrulline and Arginine Following Single-dose Supplementation with Citrulline

All the citrulline supplements examined resulted in significant increases in plasma citrulline (Figure 2) with approximately 15 to 75-fold increases over baseline levels taken prior to supplementation observed (Table 2). Peak plasma concentrations of citrulline occurred within 60 minutes of oral dosing and were greatest in the L-citrulline treatment group followed by the 6 g citrulline HCl and the 2 g citrulline HCl treatment groups (Figure 2A; Table 2). Citrulline supplementation also resulted in time-dependent increases in plasma arginine levels (Figure 2B). Despite the reduced levels of plasma citrulline observed with the 6 g citrulline HCl formulation, increases in plasma arginine concentrations were similar to those in the 6 g L-citrulline treatment group, with both peaking at 2-3-fold above baseline within 120 minutes (Figure 2B; Table 3). Subjects receiving the 2 g oral dose of citrulline HCl also showed a time-dependent increase in plasma arginine (Figure 2B). Although the resulting Cmax was significantly lower than observed for the 6 g L-citrulline and 6 g citrulline HCl (Figure 2B; Table 3), the time required to reach Cmax was substantially shorter in the 2 g citrulline HCl treatment group, with the Tmax occurring within 60-minutes of dosing compared to 120-minutes with the 6 g L-citrulline and citrulline HCl groups (Figure 2B).

**Figure 2.**
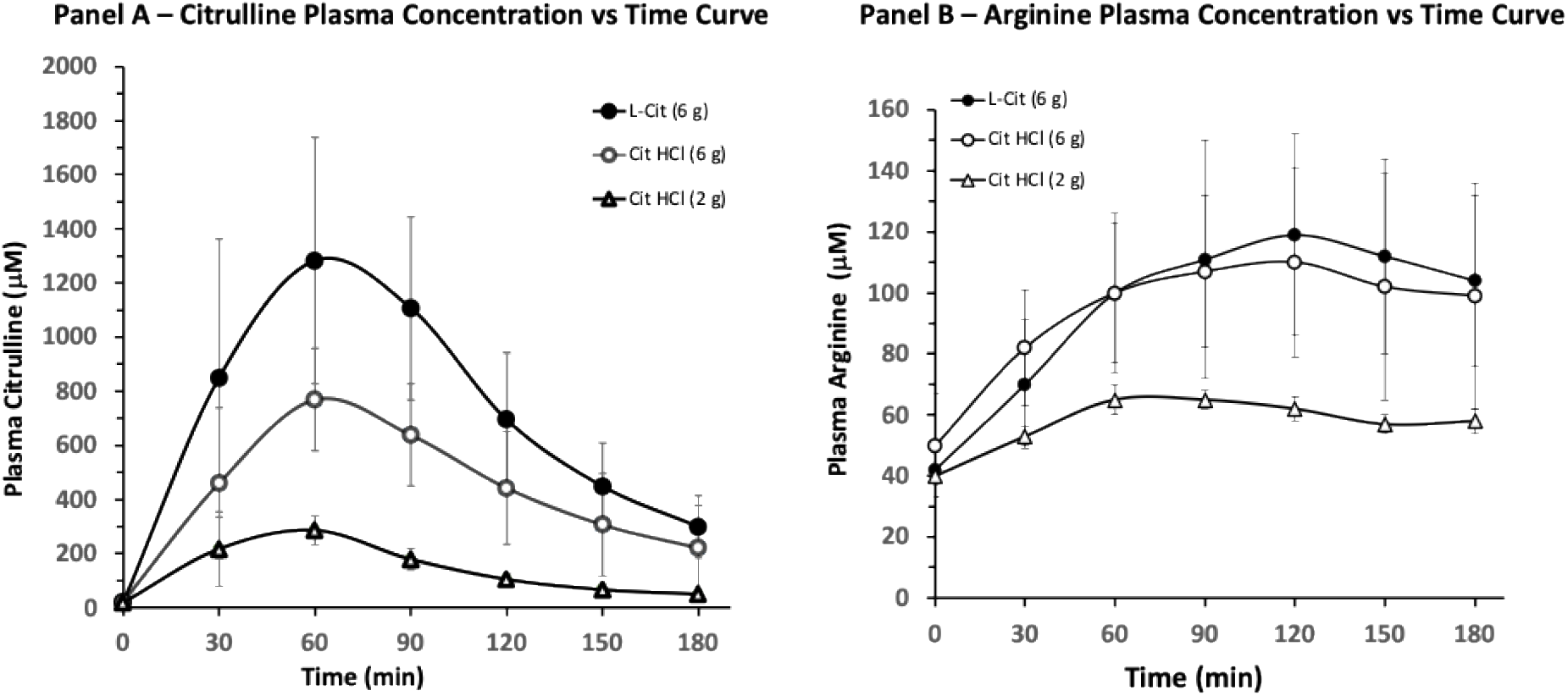
Plasma concentration vs time curves for citrulline (Panel A) and arginine (Panel B) resulting from a single bolus oral dose of citrulline. Values represent the mean ± SD of 5-6 subjects per treatment group..

**Table 2:** Citrulline plasma pharmacokinetics and renal clearance following single oral dosing with various citrulline supplements in healthy subjects.

| Treatment Group | Baseline<sup>#</sup> (μM) | C<sub>max</sub><sup>#</sup> (μM) | Increase over Baseline | Plasma AUC<sub>(0-180)</sub><sup>#</sup> (μM*min) | Plasma AUC<sub>(0-∞)</sub><sup>#</sup> (μM*min) | Citrulline CL<sub>R</sub><sup>#</sup> (ml/hr) | Fraction Citrulline<sup>#</sup> Recovered |
| --- | --- | --- | --- | --- | --- | --- | --- |
| L-citrulline 6 g | 19 ± 3 | 1421 ± 508 | 75-fold | 146501 ± 49692 | 155724 ± 50133 | 16 ± 14 | 81 ± 15 |
| Citrulline HCl 6 g | 22 ± 6 | 790 ± 198* | 36-fold | 82223 ± 24994* | 96861 ± 33187* | 13 ± 11 | 82 ± 16 |
| Citrulline HCl 2 g | 19 ± 5 | 277 ± 64** | 15-fold | 25374 ± 4625** | 29941 ± 3695** | 11 ± 11 | 88 ± 12 |

\*p<0.05 compared to L-citrulline treatment group
\*\*p<0.01 compared to L-citrulline treatment group
<sup>#</sup> Values represent the mean ± SD

**Table 3:** Arginine plasma pharmacokinetics and renal clearance following single oral dosing with various citrulline supplements in healthy subjects.

| Treatment Group | Baseline<sup>#</sup> (μM) | Cmax<sup>#</sup> (μM) | Increase over Baseline | Plasma AUC<sub>0-180</sub><sup>#</sup> (μM*min) | Plasma AUC<sub>(0-∞)</sub><sup>#</sup> (μM*min) | Arginine CL<sub>R</sub><sup>#</sup> (ml/hr) | Fraction Arginine Recovered<sup>#</sup> |
| --- | --- | --- | --- | --- | --- | --- | --- |
| L-citrulline<br>6 g | 44 ± 4 | 128 ± 35 | 2.9-fold | 19683 ±<br>4125 | 34810 ±<br>8269 | 29 ± 20 | 65 ± 25 |
| Citrulline<br>HCl 6 g | 50 ± 7 | 113 ± 31 | 2.3-fold | 17559 ±<br>4302 | 31887 ±<br>11017 | 21 ± 14 | 73 ± 15 |
| Citrulline<br>HCl 2 g | 40 ± 3 | 66 ± 15* | 1.7-fold | 10520 ±<br>1923** | 19669 ±<br>3425** | 21 ± 19 | 75 ± 18 |

\*p<0.05 compared to L-citrulline treatment group
\*\*p<0.01 compared to L-citrulline treatment group

### 3.3 Differences in Citrulline to Arginine Conversion Efficiency with Various Citrulline Formulations

As increases in plasma arginine are the result of metabolic conversion of plasma citrulline, the plasma AUCs for citrulline and arginine were used to assess the effectiveness of the various citrulline dietary supplementations in delivery of arginine (Figure 3). In the case of the 6 g Citrulline HCl treatment group, the plasma AUC_(0-∞)_ values for arginine were similar in magnitude to the 6 g L citrulline group despite a significantly lower citrulline plasma AUC_(0-∞)_ (Figure 3A and B). As expected, for the 2 g Citrulline HCl group, there were significantly lower plasma AUCs for both arginine and citrulline compared to the 6g doses of Citrulline HCl or L-citrulline (Figure 3A and B). The resulting plasma arginine to citrulline AUC ratios were used to provide an index of the conversion efficiency for each citrulline formulation (Figure 3C). Based on the resulting plasma AUC ratios for arginine and citrulline, the L-citrulline formulation was least effective in converting citrulline to arginine, with an arginine to citrulline AUC ratio of approximately 0.23 (Figure 3C). The 6 g citrulline HCl showed a moderately improved conversion ratio of 0.37 representing greater than 50% better conversion of citrulline to arginine compared to the 6 g L-citrulline formulation. However, the most efficient conversion of plasma citrulline to plasma arginine was observed with the 2 g citrulline HCl formulation with an AUC ratio of approximately 0.67 (Figure 3C), reflecting 2.8-fold better arginine conversion over the L-citrulline treatment group.

**Figure 3.**
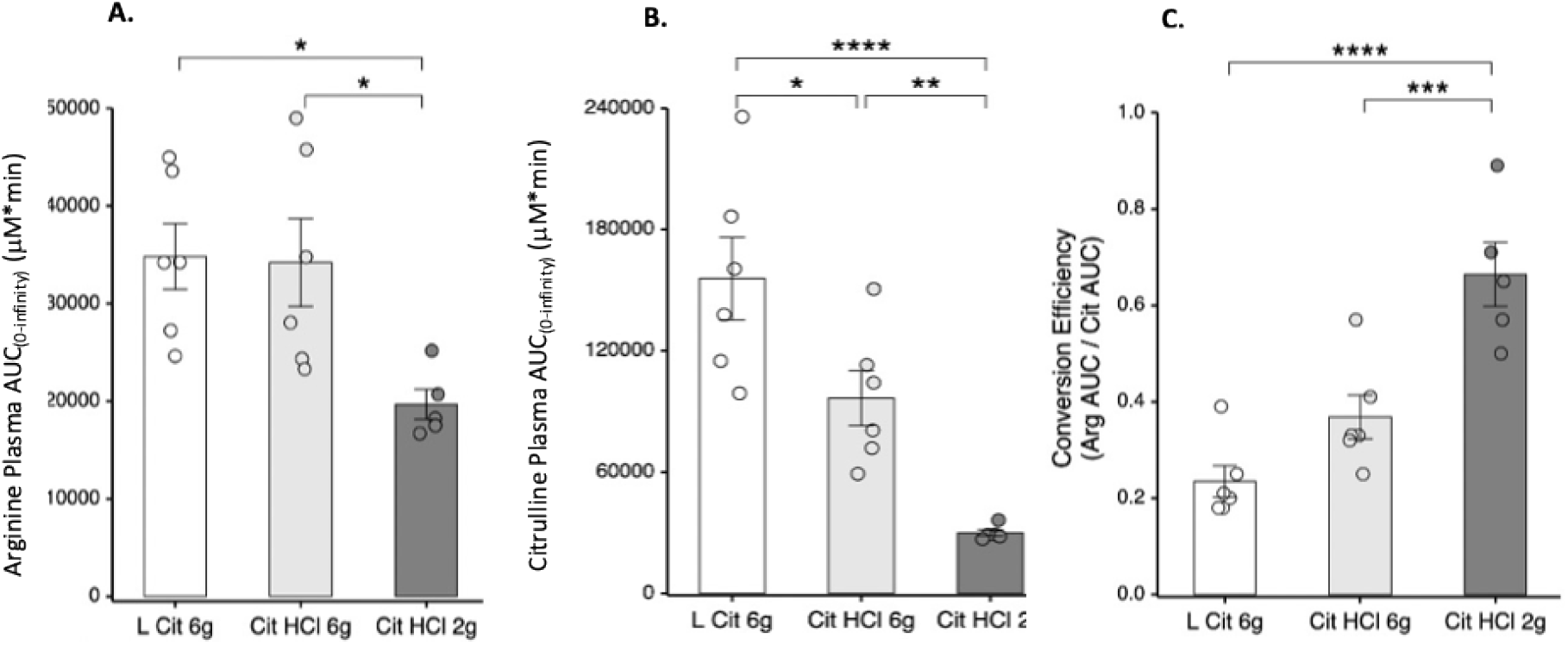
Arginine (A) and Citrulline (B) area under the plasma vs time curve and arginine conversion efficiency (C) for subjects taking 6 grams of L-citrulline or citrulline HCI at the 6 or 2 gram dosage. The bars represent the mean±SD and individual values are represented by the circles within the bar plots. * p<0.05; ** p <0.01; *** p < 0.005; **** p < 0.001 using ANOVA with Fisher post hoc comparison of the means.

### 3.4 Urinary Excretion of Arginine and Citrulline

The renal clearance rates for both citrulline and arginine were similar in all treatment groups and was reflective of amino acids with a high fraction of recovery in the kidney (Tables 2 and 3). All three citrulline supplements resulted in significant increases in 24-hr urinary output of citrulline (Figure 4A). Compared to urine citrulline levels taken prior to supplementation there was a 45-fold increase in subjects receiving L-citrulline while those receiving citrulline HCl had significantly less with only 15-fold and 7-fold increases in 24-hr urine citrulline levels observed with 6g and 2g doses, respectively (Figure 4C). While significant increases in urinary levels of arginine were also observed, they were comparable across all citrulline supplements with an approximately 5-fold increase over pre-treatment levels (Figure 4C).

**Figure 4.**
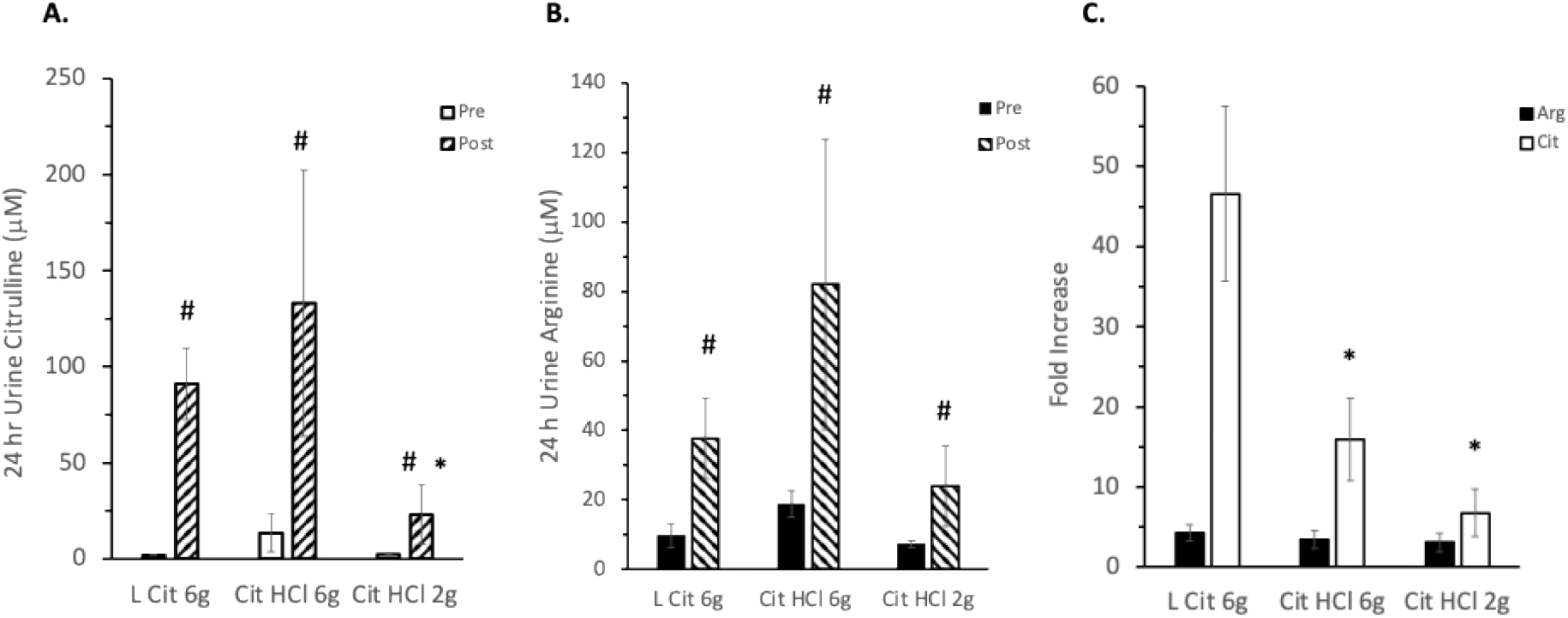
Analysis of citrulline and arginine urine concentrations following various supplement administration. Panel A-Urine citrulline concentrations over a 24-hr period prior to and immediately following single dose administration of various citrulline dietary supplements. Panel B - Urine arginine concentrations over a 24-hr period prior to and immediately following single dose administration of various citrulline dietary supplements. dietary supplements. Panel C - Fold-increases in citrulline and arginine over pre-dosing levels Values represent the mean ± SEM of 5-6 subjects per group. * p < 0.05 compared to L Cit 6g treatment group; # p<0.05 compared to 24-h pretreatment values within each treatment group.

### 3.5 Bioavailability Comparisons of the Various Citrulline Formulations

The relative bioavailability of the various dietary supplements are shown in Figure 5A. From a citrulline bioavailability standpoint, the 2 and 6 g doses of citrulline HCl had a lower bioavailability compared to L-citrulline (Figure 5A). In contrast, the relative bioavailability for arginine was greater for both the 2 g and 6 g citrulline HCl treatment groups compared to the 6 g L-citrulline formulation (Figure 5A). For the 6 g citrulline HCl formulation the relative arginine bioavailability was approximately 120% compared to 6 g L-citrulline. However, for the 2 g citrulline HCl formulation the relative arginine bioavailability was over 200% greater compared to L-citrulline (Figure 5A). These findings are consistent with the improved arginine conversion observed with citrulline HCl formulations. Indeed, when the increases in plasma arginine were normalized to the dose of citrulline administered, the 6 g citrulline HCl (representing a 4.95 g adjusted dose of citrulline) was equivalent to 6 g L-citrulline, and the 2 g citrulline HCl (representing a 1.65 g adjusted dose of citrulline) was superior to all other citrulline formulations (Figure 5B).

**Figure 5.**
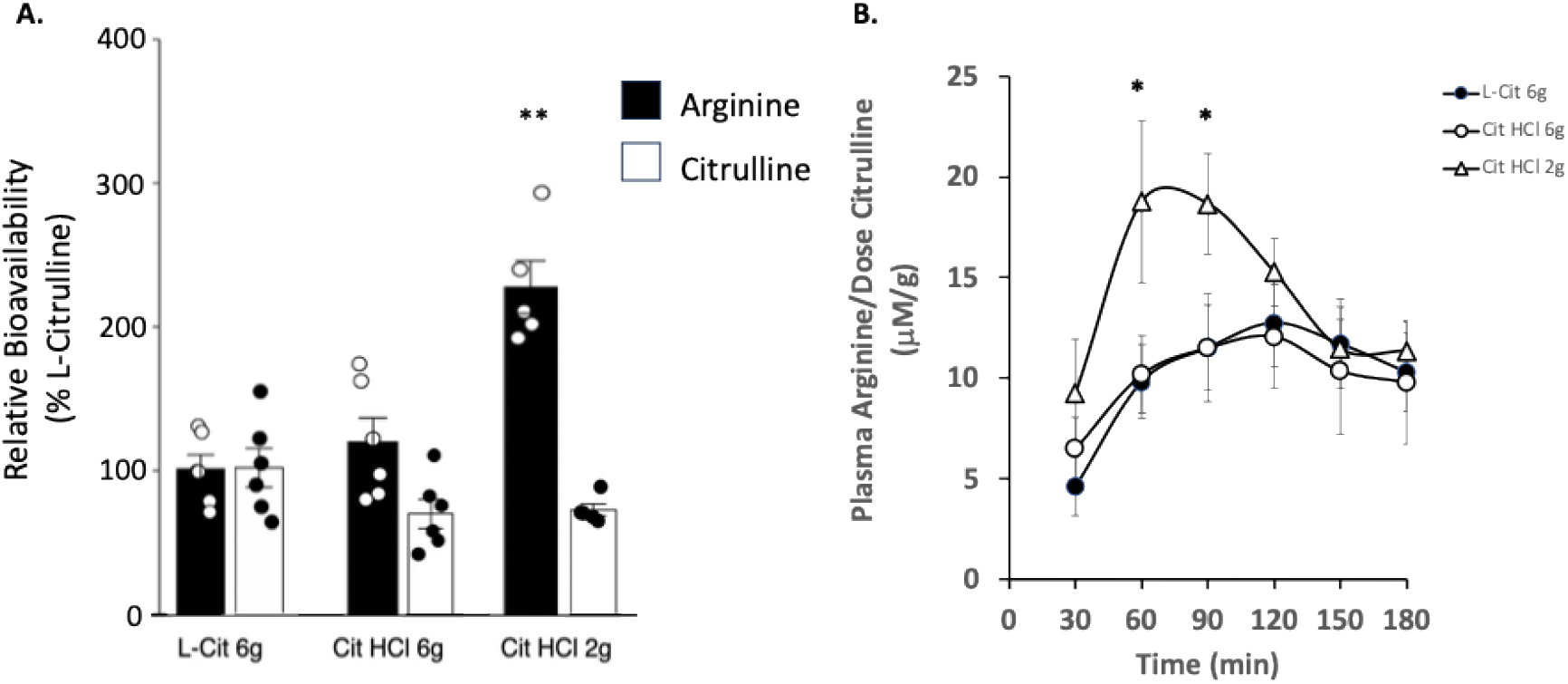
(A) Relative bioavailability of various citrulline dietary supplement formulations. Oral bioavailability of citrulline and arginine from the 6g and 2g citrulline HCI doses were compared to a 6g dose of the L-citrulline. Mean values with standard deviations are depicted in bar graph along with individual values for each group. (B) Increases in plasma arginine per dose of citrulline administered. Values represent the mean ± SEM of 5-6 subjects per treatment group. * p <0.05 compared to Cit HCI 6g and L Cit 6g treatment group at the same time point; ** p < 0.01 compared to all other citrulline formulations.

## 4. Discussion

Arginine is an important amino acid for protein production as well as providing the metabolic precursor for several molecules, such as nitric oxide and polyamines (15) that have crucial functions in health and disease. While the body can make arginine de novo, there is much interest in the health benefits derived from dietary supplementations that increase arginine levels in the blood. High dose arginine supplementation has been examined for treating a variety of pathological conditions (15-17). In addition, increasing arginine levels in the blood is believed to aid in the formation of nitric oxide within localized areas of the vasculature that lead to vasodilation and improved blood flow (2, 18, 19). While there is considerable interest in arginine supplementation, oral absorption of arginine is limited due to first pass metabolism (20, 21) and the doses of arginine required to obtain the desired therapeutic responses often have gastrointestinal side effects that limit their use (22).

Due to limitations with arginine supplementation, dietary supplementation with citrulline has emerged as a potential way to increase arginine levels in the body (23). The advantages of citrulline supplementation as a method for increasing arginine stores in the body include a better oral absorption and side-effect profile compared to arginine supplements. Dietary-sourced arginine is subject to considerable first-pass metabolism in the liver, with absolute oral bioavailability as low as 20% reported for normal healthy subjects (20). In contrast, citrulline is almost completely absorbed in the gastrointestinal tract and does not undergo first-pass metabolism. Multiple studies have reported citrulline supplementation to be equally, if not more, effective at increasing plasma arginine as supplementation with arginine (24-26). These findings, together with the reported adverse gastro-intestinal effects with arginine supplementation (22), has resulted in increased use of citrulline supplementation.

Citrulline HCl is a salt form of L-citrulline with improved formulational properties that offer potential advantages in terms of improved aqueous solubility, improved bioavailability and reduced gastrointestinal side effects. The present study compared the single-dose oral bioavailability and pharmacokinetics of citrulline HCl to a standard L-citrulline formulation and dose commonly used for dietary supplementation. All three citrulline formulations produced a rapid increase in plasma citrulline levels and displayed similar plasma kinetics as previous reported studies using L-citrulline dietary supplementation (4, 27, 28). The plasma citrulline Cmax and AUC values were greatest for the 6g L-citrulline formulation. All citrulline supplements also resulted in time-dependent increases in plasma arginine. Using relative bioavailability for comparing the different formulations and doses, showed the citrulline HCl formulations had improved arginine bioavailability compared to the L-citrulline formulation. These increases in arginine relative bioavailability (approximately 120% and 200% for 6 g and 2 g doses, respectively) observed following single dose administration could result in substantial elevations in arginine when used in a dietary supplement regiment.

The citrulline HCl formulation has a substantial improvement in aqueous solubility over the L-citrulline formulation. For oral absorption, improvements in the aqueous solubility would generally be expected to improve oral absorption rates in the intestine (29, 30). However, in addition to aqueous solubility, the intestinal permeability of the solute is also critical for oral absorption (30). In the case of citrulline, studies in various *in vitro* and pre-clinical animal models implicate the involvement of several different amino-acid transporters that are likely important for intestinal absorption (31, 32). Furthermore, human studies suggest citrulline is highly permeable through the gastrointestinal route (23, 33). Given the anticipated improvements in solubility and the intestinal permeability of citrulline, it is unlikely that the differences in plasma citrulline levels observed with the citrulline HCl formulation would be attributable to reduced intestinal absorption compared to L-citrulline.

Once citrulline enters the systemic bloodstream it is converted to arginine in the kidneys. While there are extrarenal sites capable of metabolizing citrulline to arginine (34), the majority occurs in the kidney through the intestinal-renal axis where citrulline within the intestines is absorbed into the bloodstream and converted to arginine in the proximal tubules of the kidney (35). Under normal physiological conditions, the renal excretion of citrulline and arginine is low due to reabsorption of these solutes in the proximal tubules of the kidney (35). Previous studies have reported that even with high loading doses of orally administered citrulline (ie. 15 g) the urinary excretion of citrulline was less then 5% (4). In the present study, both the renal clearance (Cl_R_Cit) and fractional recovery (F_r_Cit) of citrulline were similar in all three treatment groups and characteristic of a solute with low renal elimination. Thus, it is unlikely that altered renal clearance has a role in the differences in citrulline bioavailability observed.

Absent demonstrable differences in the intestinal absorption and renal elimination of citrulline, the remaining explanation for the lower citrulline plasma levels observed in the present study with the citrulline HCl formulations is a more efficient conversion to arginine. Multiple lines of evidence support this possibility. First, the relative bioavailability of arginine is greater in the citrulline HCl formulations compared to L-citrulline. In the case of the 2g citrulline HCl formulation there is approximately 2-fold improvement in arginine bioavailability compared to 6 g L-citrulline. Secondly, the plasma AUC ratios for the metabolite (arginine) and precursor (citrulline) are higher for the 2 g (approximately 3-fold) and 6g (approximately 1.5-fold) citrulline HCl compared to the L-citrulline treatment group. These findings support a higher (i.e. more efficient) rate of conversion of citrulline to arginine with the citrulline HCl formulations. Finally it should also be noted that the increases in urinary citrulline compared to pre-treatment baseline levels were significantly greater for 6 g L-citrulline compared to either the 6 g or 2 g citrulline HCl formulations while increases in arginine excreted in the urine were similar for all three supplements. These findings are consistent with potential saturation of the renal enzymes responsible for converting citrulline to arginine which were most apparent with the 6 g L-citrulline formulation and suggest the citrulline HCl formulations appear to have improved conversion efficiency and less excretion of citrulline in the urine. As the intent of citrulline dietary supplementation is to increase arginine available for tissues, the citrulline HCl formulation does appear to provide improved performance compared to L-citrulline.

Previous studies examining escalating doses of L-citrulline suggested that doses above 10 g were not as effective as lower doses (4). The dose-dependency observed at high doses (> 10 g) of citrulline supplementation were attributed to saturation of absorption routes in the intestine. The findings in the current study suggest that there may also be dose-dependent effects in terms of the citrulline to arginine conversion within the kidneys, and these effects may be apparent at even lower doses of citrulline supplementation. Further studies are warranted in this regard.

Although the present study is a single-dose study there are some interesting observations to suggest the citrulline HCl formulation may have advantages over L-citrulline. First, when considered on a per dose basis, 2 g citrulline HCl produced the greatest increases in plasma arginine. This is likely due to the increased rate of conversion of plasma citrulline to arginine in the kidney following dosing with 2 g citrulline HCl. Furthermore, the time required to attain peak arginine levels (Tmax) was significantly shorter with the 2 g citrulline HCl formulation. For those taking a citrulline supplement to potentially increase blood flow and protein accrual in the muscle (36, 37) a shorter lag time for achieving peak levels of arginine in the blood may be a desired property. Based on the present single-dose pharmacokinetic studies showing improved arginine conversion with the 2 g citrulline HCl formulation, there may be advantages in splitting a single daily dose into smaller multiple daily dosing units to optimize arginine delivery.

## Data Availability

All Data produced in the present study are available upon reasonable request to the authors

## Acknowledgements

Authors wish to thank Dr. Donald W. Miller (Department of Pharmacology and Therapeutics, University of Manitoba) for his help with pharmacokinetic analysis of the data presented in the manuscript. Financial support for the studies was provided by Vireo Systems, Williams, TN.

